# Knowledge and perceptions of COVID-19 among government employees in Ethiopia

**DOI:** 10.1101/2020.11.12.20230615

**Authors:** Wakgari Deressa, Alemayehu Worku, Wondwosson Amogne, Sefonias Getachew, Awgichew Kifle, Workeabeba Abebe

**Affiliations:** Department of Preventive Medicine, School of Public Health, College of Health Sciences, Addis Ababa University, Addis Ababa, Ethiopia; Department of Internal Medicine, School of Medicine, College of Health Sciences, Addis Ababa University, Addis Ababa, Ethiopia; Department of Pediatrics and Child Health, School of Medicine, College of Health Sciences, Addis Ababa University, Addis Ababa, Ethiopia

**Keywords:** COVID-19, Ethiopia, Government employees, Knowledge, Perception, SARS-COV-2

## Abstract

**Background:** In the absence of effective treatments or vaccines, the spread of the novel coronavirus disease 2019 (COVID-19) pandemic can be minimized by effectively implementing preventive measures. Knowledge and perceptions of the public about COVID-19 play a critical role in behavioral changes. This study aimed to assess the knowledge and perceptions of COVID-19 as well as source of information about the disease among government employees.

**Methods:** A cross-sectional survey of 1,573 government employees from 46 public institutions located in Addis Ababa was undertaken from 8^th^ to 19^th^ June 2020. Paper-based self-administered questionnaires were used for data collection. ANOVA test and t-test were used to assess the difference between groups.

**Results:** The respondents demonstrated very high knowledge of the cause of COVID-19 (93%), its main clinical symptoms (>90%), the main modes of transmission (89%), the main preventive measures (>90%). Almost all respondents reported that people with recent travel history (86.8%) or people who had contact with COVID-19 patients (93.5%) were the high-risk groups to be infected with coronavirus. In addition, more than half (50.9%) of the study participants reported that people without travel history nor had contact with confirmed cases are also most likely to be infected with the virus. About 84% of the respondents perceived that older adults and elderly were most at risk to die from COVID-19. Similarly, the majority of the respondents reported that adults with other underlying health problems (95.4%), cigarette smokers (88.1%) and substance users (87.5%) were more likely to die from the disease. An electronic media such as television (85.5%), social media (74.1%), online materials (71.1%) and radio (60.8%) constituted the primary sources of information about COVID-19, followed by healthcare workers (66.6%) and print materials (35.4%). Television (32.2%) and health workers (30.5%) constituted the most trusted sources of information related to COVID-19.

**Conclusions:** This study has showed higher level of knowledge and favorable perception among respondents about COVID-19. Knowledge and perceptions have great roles in behavioral change and efforts should be focused on improving the perceived susceptibility, severity, and benefits of preventive behavioral changes by providing timely and adequate information.

## Background

The current worldwide rapid spread of the novel coronavirus disease 2019 (COVID-19), caused by severe acute respiratory syndrome coronavirus 2 (SARS-CoV-2), has led the World Health Organization (WHO) to declare it as a full global pandemic on 11^th^ March 2020 [1]. This unprecedented rapid transmission of the virus has already reached all countries and territories around the world and caused over 49 million confirmed cases and more than 1.2 million deaths worldwide as of 5^th^ November 2020 [2]. As of 5^th^ November, a total of 1,854,169 COVID-19 cases and 44,316 deaths were reported from 57 countries and territories in Africa. The rapid transmission of the virus from person to person coupled with lack of effective medications and vaccines has posed serious challenges to controlling the spread of the disease.

The first confirmed COVID-19 case in Ethiopia was reported on 13^th^ March 2020. As of 5^th^ November, a total of 98,391 confirmed COVID-19 cases with over 57,000 recoveries and 1,508 deaths were reported in the country [2]. This is the highest number of COVID-19 confirmed cases in East Africa and 4^th^ across Africa. Most (83%) of the cases during the first couple of months were either imported or close contacts of the imported cases [3] and later dominated by the community transmission [4]. As of 25^th^ October, contacts of the confirmed cases contributed for about 23% of the total COVID-19 cases reported in Ethiopia [5]. During our data collection period, the number of nationally confirmed cumulative COVID-19 cases increased from 2,156 on 8^th^ June to 4,070 on 19^th^ June 2020, with 89% increase in less than two weeks. Children under the age of five years and between the ages of 5-15 years were also tested positive for COVID-19. As to the current scenario, the overall positivity and case fatality rates of Ethiopia are about 6.5% and 1.5%, respectively [5].

The initial mitigation measures of COVID-19 in Ethiopia focused on isolating and treating confirmed cases at the treatment center, tracing and quarantining contacts, 14 days mandatory quarantine of all passengers coming to the country, risk communication and educating the public about preventive measures to reduce the risk of transmission. Subsequently, Ethiopia has adopted the recommendations of WHO’s basic public health measures to reduce and contain the transmission of COVID-19 [6, 7]. In addition, the Government of Ethiopia has implemented a variety of policy actions and precautionary measures in response to the COVID-19 pandemic, such as airport surveillance and suspension of flights, travel restrictions, closure of international borders, flexible working arrangements, closing schools and universities, suspending sporting and religious gatherings. Finally, state of emergency that lasted between 11^th^ April to 10^th^ September 2020 was declared to counter and control the spread of COVID-19 and mitigate its impact [8].

Intense media coverage in Ethiopia about the COVID-19 outbreak began in early February 2020 and peaked in March after WHO declared its pandemic level [1] and the first confirmed case report in the country on 13^th^ March 2020. Since then, awareness creation and updates of the situation of the pandemic in the country have been daily given to the public by the Ministry of Health (MoH) and Ethiopian Public Health Institute (EPHI) on mass media including radio and television. In addition, daily updates of the COVID-19 situation and different risk communication and awareness creation messages have been posted on their Websites, Facebook and Twitter pages. Practically, all mass media in the country devoted extensive time to the coverage of various aspects of the pandemic of COVID-19 targeting the general public. Government health education messages via various mass media are the major source of information about COVID-19 and for promoting self-protective practices. These preventive messages generally emphasize frequently washing hands with soap and water, mask wearing, and social distancing measures, including staying at home as much as possible, avoiding close contact with people including shaking hands, and avoiding crowds and mass gatherings. However, the public uptake of such health protective behaviors during epidemics relies on the trust in media and government information [9].

Despite efforts to scale-up public health interventions to contain and mitigate the spread of coronavirus in Ethiopia, new cases have continued to emerge. The coronavirus has rapidly spread to every part of the country and local community transmission has been established. Gaps and negligence in applying the COVID-19 preventive measures among the public have been observed. The literature on the previous outbreaks of infectious disease suggests that more knowledgeable people during periods of disease outbreaks tend to worry more about being infected, suggesting a link between knowledge and risk perception [10]. Therefore, correct knowledge and positive perceptions and attitudes have paramount importance to prevent the transmission of the virus through enhancing the preventive practice of people. Studies of people’s perspectives provide timely and relevant evidence to identify the best preventive intervention that may be required to correct misconceptions and misunderstandings about the virus [11].

Although there is relatively high knowledge of COVID-19 among urban residents in Ethiopia, there are several misconceptions and understandings associated with the disease [12]. Likewise, people’s adherence and compliance to the preventive measures and law enforcement is generally weak to stop the nationwide spread of the virus. Since the first known confirmed COVID-19 case in Ethiopia, very few studies were conducted to assess knowledge and perceptions of the people as part of the efforts in the fight against the disease [13-15]. It is therefore important to understand people’s knowledge and perceptions regarding COVID-19 to develop effective messages. The aim of this study was to rapidly assess the knowledge and perceptions of COVID-19 among government employees in Addis Ababa. The results of this study are important to inform future efforts focusing on the people’s readiness to comply with pandemic control measures and the development of preventive strategies and health promotion programs.

## Methods

### Study area and setting

This study was conducted between 08-19 June 2020 in Addis Ababa city administration, three months after the first confirmed COVID-19 case was reported in Ethiopia. Addis Ababa city is administratively divided into 10 sub-cities and 116 districts. The projected population of the city was about 3.6 million in 2019, and was estimated to reach 4.5 million inhabitants by 2030 [16]. Addis Ababa city has the highest burden of COVID-19 cases and deaths in Ethiopia. As of October 18, a total of 45,1724 confirmed COVID-19 cases were reported from Addis Ababa, representing about 51% of the national cases. The responses to COVID-19 pandemic are in line with the national response strategy, focusing on physical distancing; hand washing; universal use of masks in public places; avoiding crowding and mass gatherings; tracing, testing, isolating and quarantining; closures of schools, places of worship, and sporting events; limiting mobility and stay at home approaches. However, there has been a lot of misconceptions in the city about the COVID-19 pandemic, how the virus itself spreads and the necessary precautions that should be taken to prevent infection. In addition, policy responses to COVID-19 have an impact on the socio-economic functioning of the people and are likely to impact their daily livelihoods.

### Study design and sampling

This institution-based cross-sectional paper-based self-administered survey was conducted among government employees of 46 public institutions located in Addis Ababa city administration. As it was not feasible to conduct a representative nationwide household or online survey during this period, the researchers opted to use self-administered survey. The study population for this study constituted all government employees, working in the selected government institution at the time of the survey and willing to participate in the study. A single cross-sectional study design sample size calculation was used based on a 50% prevalence estimate of the outcome variable at 95% confidence level, 4% precision, a design effect of 2 and 30% non-response rate. Accordingly, the minimum targeted total sample size for this survey was 1,560 respondents.

Purposive sampling was used to select public institutions located in Addis Ababa city administration. The institutions were mainly selected from the national or Federal Government Ministries, Addis Ababa city administration bureaus and sub-cities, and Oromia Regional State bureaus located in Addis Ababa. A total of 14 Ministry offices, 12 bureaus and six sub-city offices of Addis Ababa city administration, 12 bureaus of Oromia Regional State, a National Bank of Ethiopia, and Commercial Bank of Ethiopia Addis Ababa branch were included [Additional file 1]. The reasons for selecting those institutions were because of their diversity in terms of the types of employees, education, experience and their involvement in decision making processes at various regional and national hierarchies. Proportional quota sampling was used to allocate the sample size, with quotas based on the estimated number of employees per institution

### Data collection

The survey tools, self-administered questionnaires (SAQ), were developed by the research team. Some questions were adopted from the WHO tool for behavioral insights for COVID-19 [17]. The SAQ was initially prepared in English and the final version was then translated into *Amharic* and *Afan Oromo* by two experienced personnel in order to make the questions shorter, simpler and understandable for the respondents. The tools were again back translated into English by other independent personnel in order to ensure consistency. The questions in the SAQ were organized into five sections: (i) socio-demographics and other details; (ii) knowledge of COVID-19 signs and symptoms, (iii) knowledge of the mode of transmission, (iv) knowledge of treatment and vaccine; (iii) knowledge of prevention methods; and (iv) source of information.

The final English questionnaire was translated independently into *Amharic* and *Afan Oromo* languages. Backward translation from *Amharic* and *Afan Oromo* to English was carried out by another two independent translators. A total of 20 data collectors were involved in the data collection of this survey, using standardized paper-based SAQ. After obtaining informed consent from all potential participants, the questionnaires were distributed to the respondents. The respondents were also informed neither to consult a friend nor read an online or any other material before responding to the questions.

### Statistical analyses

Data were entered into the Census Surveys Professional (CSPro) version 7.2 statistical software package and imported into the Statistical Package for Social Sciences (SPSS) version 23 (SPSS Inc., IBM, USA) for cleaning and analysis. Most of the knowledge and perception questions were designed as ‘yes’ or ‘no’. Knowledge of the cause, mode of transmission, main clinical symptoms and preventive measures was assessed using a 15-item scales with responses recoded as ‘1=Correct’ and ‘0=Incorrect’. The items were summed to form a total knowledge score, with higher scores indicating greater knowledge (range=0-15). Cronbach’s alpha for the knowledge score was 0.786.

Basic descriptive statistical methods such as frequencies, percentages and cross-tabulations were conducted to summarize the data and determine the differences between groups for selected demographic variables. Descriptive statistics by level of government were summarized using frequency distribution tables. Knowledge scores were compared according to demographic characteristics with independent samples *t-*test for two groups, or one-way between-groups analysis of variance (ANOVA) with Tukey HSD post-hoc tests for three or more groups. Multivariable linear regression analysis was not performed to identify factors associated with knowledge of COVID-19 due to the violation of normality assumption. The statistical significance level was set at p*<*0.05.

## Results

### Characteristics of the sample

Of 1,730 eligible participants contacted from 46 public or government institutions, 1,573 (91%) successfully completed the interview for final analysis. About 40% of the respondents were drawn from national institutions, followed by Addis Ababa city administration (38.8%) and Oromia Regional State (21.6%). The mean (±SD) age of the respondents was 35 (±8.5) years (range: 19-66) and participants aged 30-49 years represented 63.9% of the sample. The majority of the respondents 64.2% were male and 88.3% held a bachelor’s degree or above. The mean (±SD) and median years of service in the institution was 6.6 (±6.4) and 4 years (range: 1-37), respectively. The mean (±SD) household size was 4.1 (±2) people (range: 1-14). Table 1 shows the distribution of the participants’ characteristics by government level. The distributions of the characteristics across the three government levels were similar to each other except that majority of respondents in Oromia resided out of Addis Ababa (33.3%) and had at least 10 years of service (48.9%).

**Table 1.** Characteristics of study participants by level of government

| Characteristics | Government level, n (%) |  |  | Total, n (%) |
| --- | --- | --- | --- | --- |
|  | National | Addis Ababa | Oromia |  |
| **Gender (n=1,532)** |  |  |  |  |
| Male | 405 (66.3) | 350 (58.9) | 228 (69.7) | 983 (64.2) |
| Female | 206 (33.7) | 244 (41.1) | 99 (30.3) | 549 (35.8) |
| **Age group (years) (n=1,436)** |  |  |  |  |
| 18-29 | 174 (30.4) | 175 (31.3) | 48 (15.7) | 397 (27.6) |
| 30-39 | 253 (44.2) | 258 (46.2) | 145 (47.5) | 656 (45.7) |
| 40-49 | 90 (15.7) | 94 (16.8) | 77 (25.2) | 261 (18.2) |
| ≥50 | 55 (9.6) | 32 (5.7) | 35 (11.5) | 122 (8.5) |
| **Level of education (n=1,533)** |  |  |  |  |
| ≤12<sup>th</sup> grade | 17 (2.8) | 23 (3.8) | 7 (2.2) | 47 (3.1) |
| Diploma* | 53 (8.7) | 65 (10.7) | 14 (4.4) | 132 (8.6) |
| Bachelor's degree | 318 (52.3) | 391 (64.6) | 180 (56.3) | 889 (58.0) |
| Master's degree or above | 220 (36.2) | 126 (20.8) | 119 (37.2) | 465 (30.3) |
| **Year of experience in the institution (n=1,517)** |  |  |  |  |
| <5 | 352 (58.0) | 228 (55.1) | 82 (26.0) | 762 (50.2) |
| 5-9 | 160 (26.4) | 167 (28.1) | 79 (25.1) | 406 (26.8) |
| 10-14 | 44 (7.2) | 61 (10.3) | 76 (24.1) | 181 (11.9) |
| ≥15 | 51 (8.4) | 39 (6.6) | 78 (24.8) | 168 (11.1) |
| **Household size (n=1,506)** |  |  |  |  |
| 1-3 | 267 (44.6) | 227 (38.2) | 89 (28.4) | 583 (38.4) |
| 4-5 | 231 (38.6) | 249 (41.8) | 142 (45.4) | 622 (41.3) |
| 6-7 | 76 (12.7) | 85 (14.3) | 65 (20.8) | 226 (15.0) |
| ≥8 | 24 (4.0) | 34 (5.7) | 17 (5.4) | 75 (5.0) |
| **Area of residence (n=1,502)** |  |  |  |  |
| Addis Ababa city | 580 (94.0) | 584 (95.9) | 129 (46.7) | 1,297 (86.1) |
| Out of Addis Ababa | 37 (6.0) | 25 (4.1) | 147 (53.3) | 209 (13.9) |

\*12<sup>th</sup> grade complete and 1 or more years of training

### Knowledge of the causes, signs and symptoms, incubation period and treatment of COVID-19

The majority (92.9%) of the study participants knew that COVID-19 was caused by a viral infection, and this was consistently higher (>91%) across the three government levels (Table 2). Meanwhile 89.2% of the respondents correctly identified the mode of SARS-COV-2 transmission via respiratory droplets produced when an infected person coughs or sneezes. About 4% of the respondents stated that the coronavirus can be transmitted via airborne droplets or through eating uncooked food. The most commonly cited signs and symptoms of the COVID-19 included fever (96.9%), cough (95.2%), difficulty in breathing (95.2%), headache (90.3%), sore throat (83.9%) and myalgia (81%). The less commonly reported signs and symptoms comprised loss of taste and smell, runny nose, diarrhea and hemoptysis (coughing up of blood). About 62% of the respondents from Oromia reported runny nose as a symptom of COVID-19 as opposed to 43.6% and 40% of respondents from national or Addis Ababa, respectively. Nearly 93% of the respondents cited correctly that the maximum incubation period for the novel coronavirus was up to 14 days. About 95% of the respondents correctly stated the unavailability of specific vaccine or antiviral treatment for COVID-19. Few respondents cited the availability of specific vaccine or treatment for COVID-19.

**Table 2.** Distribution of novel coronavirus related knowledge of study participants by level of government

| Knowledge | Government level, n (%) |  |  | Total, n (%) |
| --- | --- | --- | --- | --- |
|  | National | Addis Ababa | Oromia |  |
| Cause of COVID-19 infection |  |  |  |  |
| Virus | 578 (92.6) | 558 (91.5) | 326 (96.2) | 1,462 (92.9) |
| Bacteria | 15 (2.4) | 23 (3.8) | 4 (1.2) | 42 (2.7) |
| Parasite | 10 (1.6) | 8 (1.3) | 4 (1.2) | 22 (1.4) |
| Air pollution or climate change | 5 (0.8) | 7 (1.1) | 1 (0.3) | 13 (0.8) |
| Contaminated food or water | 5 (0.8) | 3 (0.5) | 1 (0.3) | 9 (0.6) |
| Other | 11 (1.8) | 11 (1.8) | 3 (0.9) | 25 (1.6) |
| Modes of transmission |  |  |  |  |
| Spread through respiratory droplet | 562 (90.1) | 554 (90.8) | 287 (84.7) | 1,403 (89.2) |
| Spread through airborne droplet | 11 (1.8) | 19 (3.1) | 2 (0.6) | 32 (2.0) |
| Spread through eating uncooked food or raw meat | 12 (1.9) | 8 (1.3) | 12 (3.5) | 32 (2.0) |
| Other* | 39 (6.3) | 29 (4.8) | 38 (11.2) | 106 (6.7) |
| Signs and symptoms |  |  |  |  |
| Fever | 607 (97.3) | 592 (97.0) | 325 (95.9) | 1,524 (96.9) |
| Cough | 599 (96.0) | 583 (95.6) | 315 (92.6) | 1,497 (95.2) |
| Shortness of breathing | 593 (95.0) | 583 (95.6) | 321 (94.7) | 1,497 (95.2) |
| Sore throat | 534 (85.6) | 492 (80.7) | 294 (86.7) | 1,320 (83.9) |
| Headache | 562 (90.1) | 549 (90.0) | 309 (91.2) | 1,420 (90.3) |
| Myalgia or fatigue | 511 (81.9) | 478 (78.4) | 285 (84.1) | 1,274 (81.0) |
| Coughing blood | 194 (31.1) | 165 (27.0) | 140 (41.3) | 499 (31.7) |
| Runny nose | 272 (43.6) | 244 (40.0) | 209 (61.7) | 725 (46.1) |
| Diarrhea | 277 (44.4) | 232 (38.0) | 139 (41.0) | 648 (41.2) |
| Loss of taste and smell | 315 (50.5) | 250 (41.0) | 209 (61.7) | 774 (49.2) |
| Other | 25 (4.0) | 27 (4.4) | 13 (3.8) | 65 (4.1) |
| Maximum incubation period |  |  |  |  |
| Up to 3 days | 2 (0.3) | 6 (1.0) | 5 (1.5) | 13 (0.8) |
| Up to 7 days | 19 (3.0) | 26 (4.3) | 15 (4.4) | 60 (3.8) |
| Up to 14 days | 588 (94.2) | 562 (92.1) | 310 (91.4) | 1,460 (93.4) |
| Don't know | 15 (2.4) | 16 (2.6) | 9 (2.7) | 40 (2.5) |
| Treatment or vaccine |  |  |  |  |
| There is currently neither treatment or vaccine | 600 (96.2) | 585 (95.9) | 312 (92.0) | 1,497 (95.2) |
| Specific treatment available | 4 (0.6) | 6 (1.0) | 1 (0.3) | 11 (0.7) |
| Specific vaccine available | 3 (0.5) | 4 (0.7) | 5 (1.5) | 12 (0.8) |
| Both treatment and vaccine available | 12 (1.9) | 12 (2.0) | 10 (2.9) | 34 (2.2) |
| Don't know | 5 (0.8) | 3 (0.5) | 11 (3.3) | 19 (1.2) |
| Total, n (%) | 624 (100) | 610 (100) | 339 (100) | 1,573 (100) |

\*Includes hand shaking, physical contact with people or covid-19 patient, contaminated water or food

### Knowledge of COVID-19 preventive measures

Respondents’ knowledge level of coronavirus virus prevention methods across the government level was pervasive (Table 3). The overwhelming majority (>90%) of the respondents reported very high knowledge of coronavirus prevention that includes the most important prevention methods such as stay at home, physical distancing, avoiding close contact with people including hand shaking, use of face mask, frequent hand washing with water and soap, avoiding touching eyes, nose and mouth, avoiding mass gatherings, movement restriction and use of sanitizers/disinfectants. Of the respondents, only 34.6 % thought that they had very good knowledge on how to prevent the spread of novel coronavirus, and 56.5% reported good or adequate knowledge to prevent the spread of the virus.

**Table 3.** Distribution of respondents’ knowledge of coronavirus preventive measures

| Knowledge | Government level, n (%) |  |  | Total, n (%) |
| --- | --- | --- | --- | --- |
|  | National | Addis Ababa | Oromia |  |
| Knowledge of novel coronavirus prevention |  |  |  |  |
| Staying at home | 598 (95.8) | 589 (96.6) | 317 (93.5) | 1,504 (95.6) |
| Physical distancing | 598 (95.8) | 587 (96.2) | 310 (91.4) | 1,495 (95.0) |
| Avoiding close contact with people including hand shaking | 599 (96.0) | 574 (94.1) | 314 (92.6) | 1,487 (94.5) |
| Use of face mask | 603 (96.6) | 574 (94.1) | 317 (93.5) | 1,494 (95.0) |
| Frequent hand washing with water and soap | 600 (96.0) | 577 (94.6) | 316 (93.2) | 1,493 (94.9) |
| Avoiding touching eyes, nose and mouth | 600 (96.2) | 567 (93.0) | 309 (91.2) | 1,476 (93.8) |
| Avoiding mass gatherings | 589 (94.4) | 565 (92.6) | 315 (92.9) | 1,469 (93.4) |
| Movement restriction | 567 (90.9) | 550 (90.2) | 312 (92.0) | 1,429 (90.8) |
| Use of sanitizers/disinfectants | 569 (91.2) | 547 (89.2) | 301 (88.8) | 1,417 (90.1) |
| Other | 30 (4.8) | 30 (4.9) | 16 (4.7) | 78 (5.0) |
| Self-rated knowledge on how to prevent the spread of novel coronavirus |  |  |  |  |
| Very good knowledge | 229 (36.7) | 226 (37.0) | 90 (26.5) | 545 (34.6) |
| Good knowledge | 350 (56.1) | 337 (55.2) | 201 (59.3) | 888 (56.5) |
| Poor knowledge | 31 (5.0) | 33 (5.4) | 33 (9.7) | 97 (6.2) |
| Very poor knowledge | 9 (1.4) | 12 (2.0) | 9 (2.7) | 30 (1.9) |
| Don't know | 5 (0.8) | 2 (0.3) | 6 (1.8) | 13 (0.8) |
| Total, n (%) | 624 (100) | 610 (100) | 339 (100) | 1,573 (100) |

### Perceptions about susceptibility and severity of COVID-19

Almost all respondents correctly stated that people with recent travel history abroad (86.8%) or people with contact with COVID-19 patients (93.5%) were the high-risk group most likely to be infected with the virus (Table 4). However, more than half (50.9%) of the study participants reported that people without travel history nor had contact with confirmed COVID-19 cases are also most likely to be infected with coronavirus. About 84% of the respondents perceived that older adults and elderly (above 60 years) were most at risk to die from COVID-19. Meanwhile, 62.8% of the study participants reported that both male and female have equal chance to die from the disease although 22.2% perceived that males had more chance to die from COVID-19. With the group of adults more likely to die if contracted COVID-19, the majority of the respondents reported that adults with other underlying health problems (95.4%), adults who are cigarette smokers (88.1%) and adults who frequently use substances such as alcohol, *khat* and cannabis (87.5%) were more likely to die from COVID-19. In contrast, just over one-fifth (21.9%) of the respondents reported that adults without other underlying health problems are also most likely to die from COVID-19.

**Table 4.** Distribution of responses on perceptions about susceptibility and severity of COVID-19

| Susceptibility and severity perceptions | Government level, n (%) |  |  | Total, n (%) |
| --- | --- | --- | --- | --- |
|  | National | Addis Ababa | Oromia |  |
| **Which groups of people are most likely infected by novel coronavirus?** |  |  |  |  |
| People with recent travel history abroad | 551 (88.3) | 533 (87.4) | 281 (82.9) | 1,365 (86.8) |
| People with contact with COVID-19 patients | 583 (93.4) | 578 (94.8) | 310 (91.4) | 1,471 (93.5) |
| People with neither travel history nor contact with confirmed COVID-19 cases | 339 (54.3) | 336 (55.1) | 125 (36.9) | 800 (50.9) |
| Other | 42 (6.7) | 36 (5.9) | 21 (6.2) | 98 (6.2) |
| **Which groups of people are most likely to die from COVID-19?** |  |  |  |  |
| Children/adolescents | 4 (0.6) | 3 (0.5) | 13 (3.8) | 20 (1.3) |
| Young adults | 11 (1.8) | 9 (1.5) | 25 (7.4) | 45 (2.9) |
| Older adults/ elderly (above 60 years) | 542 (86.9) | 488 (80.0) | 286 (84.4) | 1,316 (83.7) |
| Other | 67 (10.7) | 110 (18.0) | 15 (4.5) | 192 (12.0) |
| **Is it male or female that is most likely to die from COVID-19?** |  |  |  |  |
| Male | 139 (22.3) | 125 (20.5) | 85 (25.1) | 349 (22.2) |
| Female | 16 (2.6) | 19 (3.1) | 30 (8.8) | 65 (4.1) |
| Both are equally likely to die | 413 (66.2) | 400 (65.6) | 175 (51.6) | 988 (62.8) |
| Don't know | 56 (9.0) | 66 (10.8) | 49 (14.5) | 171 (10.9) |
| **Which groups of the adults are more likely to die from COVID-19?** |  |  |  |  |
| Adults with other underlying health problems | 606 (97.1) | 578 (94.8) | 316 (93.2) | 1,500 (95.4) |
| Adults who are cigarette smokers | 565 (90.5) | 524 (85.9) | 297 (87.6) | 1,386 (88.1) |
| Adults who frequently use substances (alcohol, khat, cannabis) | 563 (90.2) | 535 (87.7) | 279 (82.3) | 1,377 (87.5) |
| Adults without other underlying health problems | 133 (21.3) | 152 (24.9) | 59 (17.4) | 344 (21.9) |
| Other | 19 (3.0) | 17 (2.8) | 12 (3.5) | 48 (3.0) |
| **Total, n (%)** | **624 (100)** | **610 (100)** | **339 (100)** | **1,573 (100)** |

### Overall knowledge score

Table 5 shows the distribution of the responses of the respondents for knowledge. The respondents demonstrated very high knowledge on the main clinical symptoms of COVID-19 (>90%), the main preventive measures (>90%), the lack of effective cure for COVID-19 (95%) and the maximum incubation period (93%).

**Table 5.** Distribution of responses of COVID-19 knowledge questions

| **Knowledge questions (n=15)** | **Correct response, n (%)** | **Other response,\* n (%)** |
| --- | --- | --- |
| What causes COVID-19 infection? | 1,462 (92.9) | 111 (7.1) |
| What is the main mode of transmission of coronavirus? | 1,403 (89.2) | 170 (10.8) |
| What is the maximum incubation period? | 1,460 (92.8) | 113 (7.2) |
| Currently there is neither treatment nor vaccine for COVID-19 | 1,497 (95.2) | 76 (4.8) |
| The main clinical symptom (fever) | 1,524 (96.9) | 49 (3.1) |
| The main clinical symptom (cough) | 1,497 (95.2) | 76 (4.8) |
| The main clinical symptom (shortness of breath) | 1,497 (95.2) | 76 (4.8) |
| The main clinical symptom (headache) | 1,420 (90.3) | 153 (9.7) |
| The main preventive measure (staying at home) | 1,504 (95.6) | 69 (4.4) |
| The main preventive measure (physical distancing) | 1,495 (95.0) | 78 (5.0) |
| The main preventive measure (avoiding close contact) | 1,487 (94.5) | 86 (5.5) |
| The main preventive measure (use of facemask) | 1,487 (94.5) | 86 (5.5) |
| The main preventive measure (frequent hand washing) | 1,493 (94.9) | 80 (5.1) |
| The main preventive measure (avoiding mass gathering) | 1,469 (93.4) | 104 (6.6) |
| The main preventive measure (use of disinfectants/sanitizer) | 1,417 (90.1) | 156 (9.9) |

\*Includes “No”, ‘Not sure’, ‘Don’t know questions’ and ‘Missing’

Table 6 shows the distribution of socio-demographic characteristics and the mean knowledge score. Overall, the majority (82.4%) of the respondents had ‘*high’* knowledge about the COVID-19. Generally, the respondents demonstrated higher knowledge scores with the mean score of 14.1±1.69, and 59% scored 100%, on a scale of 15. There was no statistically significant difference in knowledge scores between male (14.1±1.71) and female (14.0±1.92) respondents, *t* (1530) = 0.302 (two-tailed), with very small magnitude of the differences in the means (mean difference = 0.098, 95% CI: -0.088 to 0.285). Surprisingly, the difference in knowledge score is also not significant between educational groups, age groups, year of experience, household size and whether tested for coronavirus or not. In the contrary, the level of government shows statistically significant difference in knowledge score, with respondents from the Oromia less knowledgeable about COVID-19 than study participants from national or Addis Ababa, *F* (2, 1570) = 3.60, *p*=0.028.

**Table 6.** ANOVA/ t-tests for mean knowledge scores regarding COVID-19 between groups (n=15 items)

| Characteristics | Mean±SD | ANOVA/t-test | P-value |
| --- | --- | --- | --- |
| **Gender** |  |  |  |
| Male | 14.1±1.71 | 1.03 | 0.302 |
| Female | 14.0±1.92 |  |  |
| **Age group (year)** |  |  |  |
| 18-29 | 14.0±1.87 |  |  |
| 30-39 | 14.1±1.71 | 1.54 | 0.203 |
| 40-49 | 14.2±1.42 |  |  |
| 50-66 | 14.3±1.36 |  |  |
| **Education** |  |  |  |
| ≤12<sup>th</sup> grade | 13.6±2.30 |  |  |
| Diploma | 14.0±1.80 | 1.75 | 0.155 |
| Bachelor's degree | 14.1±1.85 |  |  |
| ≥Master's degree | 14.2±1.62 |  |  |
| **Experience (year)** |  |  |  |
| <5 | 14.1±1.66 |  |  |
| 5-9 | 14.0±2.02 | 0.62 | 0.599 |
| 10-14 | 14.1±1.66 |  |  |
| ≥15 | 14.2±1.45 |  |  |
| **Household size** |  |  |  |
| 1-3 | 14.0±1.99 |  |  |
| 4-5 | 14.2±1.54 | 2.37 | 0.068 |
| 6-7 | 14.2±1.43 |  |  |
| ≥8 | 14.1±1.87 |  |  |
| **Government level** |  |  |  |
| National | 14.2±1.61 |  |  |
| Addis Ababa | 14.1±1.72 | 3.60 | 0.028* |
| Oromia | 13.8±2.23 |  |  |
| **Residence** |  |  |  |
| Addis Ababa | 14.1±1.71 | 0.98 | 0.330 |
| Out of Addis Ababa | 14.0±2.16 |  |  |
| **Tested for coronavirus** |  |  |  |
| Yes | 13.9±1.96 | -1.89 | 0.068 |
| No | 14.2±1.62 |  |  |
| **Self-reported chronic illness** |  |  |  |
| Yes | 14.2±1.62 | 0.62 | 0.534 |
| No or DK | 14.1±1.71 |  |  |

\*Significant at 0.05 level (2-tailed).

### Sources of information about COVID-19

The relative importance of various sources of information about COVID-19 was assessed in this study, and the electronic media such as television (85.5%), social media (74.1%), online materials (71.1%) and radio (60.8%) constituted the primary source of information, followed by health workers (66.6%), friends/family members (54.1%) and print materials (35.4%) (Table 7). Television (32.2%) and health workers (30.5%) constituted the most trusted sources of information related to COVID-19 and the least trusted sources of information were friend/family members (0.7%) and print materials (2%). The respondents also reported a combination of trusted sources of information on COVID-19 such as radio/television, television/health workers, radio/television/health workers, websites such as MoH/EPHI and WHO.

**Table 7.** Distribution of the sources of information about COVID-19 by level of government

| Source of information | Government level, n (%) |  |  | Total, n (%) |
| --- | --- | --- | --- | --- |
|  | National | Addis Ababa | Oromia |  |
| Main sources of information about COVID-19 |  |  |  |  |
| Television | 525 (84.1) | 523 (85.7) | 297 (87.6) | 1,345 (85.5) |
| Radio | 367 (58.8) | 389 (63.8) | 200 (59.0) | 956 (60.8) |
| Print materials | 224 (35.9) | 203 (33.3) | 130 (38.3) | 557 (35.4) |
| Online materials | 453 (72.6) | 407 (66.7) | 258 (76.1) | 1,118 (71.1) |
| Social media | 473 (75.8) | 431 (70.7) | 261 (77.0) | 1,165 (74.1) |
| Health workers | 402 (64.4) | 389 (63.8) | 257 (75.8) | 1,048 (66.6) |
| Friends/family members | 328 (52.6) | 312 (51.1) | 211 (62.2) | 851 (54.1) |
| Other | 15 (2.4) | 9 (1.5) | 9 (2.7) | 33 (2.1) |
| Most trusted source of information about COVID-19 |  |  |  |  |
| Television | 209 (33.5) | 206 (33.8) | 91 (26.8) | 506 (32.2) |
| Radio | 23 (3.7) | 25 (4.1) | 15 (4.4) | 63 (4.0) |
| Print materials | 21 (3.4) | 11 (1.8) | 0.0 | 32 (2.0) |
| Online materials | 56 (9.0) | 32 (5.2) | 23 (6.8) | 111 (7.1) |
| Social media | 58 (9.3) | 65 (10.7) | 14 (4.1) | 137 (8.7) |
| Health workers | 164 (26.3) | 192 (31.5) | 123 (36.3) | 479 (30.5) |
| Friends/family members | 3 (0.5) | 8 (1.3) | 0.0 | 11 (0.7) |
| Other | 74 (11.9) | 62 (10.2) | 64 (18.9) | 200 (12.7) |
| Total, n (%) | 624 (100) | 610 (100) | 339 (100) | 1,573 (100) |

## Discussion

This study aimed to investigate the knowledge and perceptions of COVID-19 among government employees selected from 46 public institutions in Addis Ababa city administration during June 08 to 19, 2020. At the time of this study, the spread of COVID-19 in Addis Ababa cumulatively increased by 84% from 1,625 on 8^th^ June to 2,988 on 19^th^ June, with an average of 114 per day. This study revealed higher perceived knowledge about COVID-19 and its preventive measures. The prevention and control of such pandemic is strongly influenced by people’s ability to comply with the recommended preventive public health strategies, which are often associated with how people perceive their susceptibility to infection, whether they have the necessary knowledge, capacity, confidence and trust to adapt the strategies [18-22].

Given the limited epidemiological and community behavioral COVID-19 studies published in Ethiopia, we have opted to compare our findings with other studies conducted elsewhere in Africa or around the world. The respondents demonstrated sound knowledge about the cause of COVID-19, the main modes of transmission of the virus, main signs and symptoms, incubation period and unavailability of specific treatment or vaccine. Studies indicate that the most prevalent clinical symptoms among COVID-19 infected individuals include fever, cough, and fatigue [23]. Studies conducted in Ethiopia towards the end of March 2020 showed a high knowledge level about the main clinical symptoms, route of transmission of COVID-19 and preventive practices [13, 15]. Studies revealed that increased in knowledge and perceived risk to a particular health problem would influence preventive behaviors to reduce the risk of developing the health problem. This means people’s knowledge, perceptions and attitudes about COVID-19 play a central role in the prevention and control of the pandemic through influencing and shaping their specific preventive behaviors such as hygiene and social distancing [11, 24, 25]. Higher knowledge and risk perceptions about COVID-19 were associated with increased protective behavior change [26].

The current study shows that respondents were highly knowledgeable about the preventive measures of the COVID-19, with the most commonly cited measures constituting staying at home, physical distancing, avoiding close contact with people including hand shaking, use of facemask, frequently washing hands, avoiding mass gatherings, movement restriction, and use of sanitizers/disinfectants. A similar study conducted in Ethiopia in April 2020 also showed that about 90% of the participants had good prevention knowledge of maintaining physical distance and washing hands frequently [14]. Several studies conducted in other countries have also indicated high levels of COVID-19 knowledge among the general population [27-30]. The study resulted that the overall knowledge about the causes and symptoms, the unavailability of vaccine or specific antiviral treatment, and the main preventive measures of COVID-19 was ‘*high*’. This finding was not surprising since the study was conducted three months after the first confirmed COVID-19 case in Ethiopia where massive awareness creation, intensive public health interventions and policy measures have been implemented. The seriousness of the COVID-19 and the overwhelming information about the disease has helped the population to know more about this pandemic. Similar studies in Africa such as Ghana [29], Nigeria [31], Tanzania [32] and beyond [28, 30] revealed high COVID-19 knowledge among their study participants.

The findings of the current study also show that the respondents were highly knowledgeable about the group of people most at risk of contracting the novel coronavirus, and an overwhelming majority reported that people who had contact with COVID-19 patients and people with recent history of international travel were most likely for contracting the virus. These findings are associated with the number of people infected with the virus during the first couple of months after the onset of the epidemic in Ethiopia where the majority of the cases were imported from abroad or the infections were linked to the other confirmed COVID-19 cases. More than half of all respondents cited that people without a history of travel or contact with confirmed cases are also likely to acquire coronavirus infection, which is highly aligned with the current COVID-19 situation in the country, particularly with the widespread occurrence of community transmission. Current observations in Ethiopia suggest that people of all ages are generally susceptible to contract coronavirus infection. However, studies in China during the early stage of the outbreak showed that people who were in close contact with patients with symptomatic and asymptomatic COVID-19, including health care workers and other patients in the hospital, were at higher risk of infection [33].

Regarding the group of people most at risk to death after being infected by coronavirus, the majority of the study participants reported adults with other underlying health problems, older adults/ elderly, adults who are cigarette smokers and people who frequently use substances such as alcohol, *khat* and cannabis. A study from Jimma showed that 73% of the respondents knew that elderly people who have chronic illnesses are at higher risk of developing a severe form of COVID-19 [15]. A recent multivariable analysis that uses data from China finds the increasing odds of inpatient death among the elderly [34]. Another study also finds that older age and a high number of comorbidities are associated with higher severity and mortality from COVID-19 [35]. Likewise, being under 60 years with a body mass index (BMI) ranging from 30 to 34 is associated with a likelihood of being admitted to acute and critical care, compared to those with a BMI lower than 30 [36]. This reveals that a younger person who is experiencing other health complications or with higher BMI faces as much risk as the elderly. Of the COVID-19 patients investigated in another study in China, 48% suffered comorbidity, marked by a high prevalence of hypertension, diabetes, and coronary heart disease [34].

Our findings indicate that the majority of the study participants reported that both male and female have equal chance to die from COVID-19 although about one-fourth of the respondents perceived that males had more chance to die from COVID-19. Men and women generally seem to respond differently to the effects of COVID-19. Available evidence suggests that men are more likely to die of COVID-19 than women, owing presumably to the biological and lifestyle factors [37-39]. Biological factors lead to sex-differentiated susceptibility to illness. For example, men are more prone to acute health problems, partly explaining why women live a little longer than men [40]. There is also a sex difference in lifestyle or health behavior. Men smoke and consume alcoholic beverages more than women [41]. As far as the current evidence is concerned, men and women experience similar odds of COVID-19 infection, but men are disproportionately exposed to the severest of outcomes, including death [35].

During an emerging infectious disease like coronavirus, receiving timely and accurate information enables individuals to take appropriate actions to protect themselves, shaping their risk perception. In the current study television, social media, online materials, health workers and radio were identified as the main sources of information for COVID-19. In addition, television and health workers were reported as the most trusted sources of information related to COVID-19. Similar surveys conducted at the early phase of the pandemic in Ethiopia also showed social media and television to be the main source of information about COVID-19 [13,14]. In Nigeria, the most common sources of information about COVID-19 were identified as television and radio, followed by social media and the internet [42]. Under health emergency circumstances, adequate literature or reading materials may not be readily available, and in such situation, television, radio, internet and social media play a major role in providing up-to-date information to the public. However, it should be noted that access to television and social media depends on the local contexts such as on the availability of electricity and internet.

Despite generating highly valuable and timely findings, this study was not without some limitations. First, due to limited internet access and online information sources in the country, the study was based on paper-based self-administered survey that only included government employees in Addis Ababa, where vulnerable populations and illiterate people were not captured in the study. Ideally, a randomly selected sample of the public should be interviewed about COVID-19. Second, due to the threat of pandemic and the social distancing rule, it was not possible to conduct a face-to-face interview. Third, the data presented in this study are self-reported and partly dependent on the respondent’s honesty and recall ability, thus they may be subject to recall and social desirability biases. They also could have been affected by selection bias. Nevertheless, this study provides valuable information about the knowledge and perception of COVID-19 among government employees, and the findings can be used to direct specific public health interventions or messages to the government employees.

## Conclusions

The results of this study identified higher knowledge among respondents about COVID-19, including its mode of transmission, symptoms, incubation period, vulnerable population and preventive measures. Knowledge and perceptions have great roles in behavioral change and all efforts should be focused on improving the perceived susceptibility, severity, and benefits of preventive behavioral changes. Understanding how the public responds to the global pandemic like COVID-19 is useful in identifying ways of encouraging behavior change during the early stages of any infectious disease outbreak.

## Data Availability

The datasets used and analyzed during the current study are available from the corresponding author on reasonable request.

## Abbreviations

ANOVA: Analysis of variance;
AAU: Addis Ababa University;
BMI: Body mass index;
COVID-19: Coronavirus disease 2019;
CSPro: Census Surveys Professional;
EPHI: Ethiopian Public Health Institute;
HSD: Honestly significant difference;
IRB: Institutional Review Board;
MoH: Ministry of Health;
SAQ: Self-administered questionnaire;
SARS-CoV-2: Severe acute respiratory syndrome coronavirus 2;
SD: Standard deviation;
SPH: School of Public Health;
SPSS: Statistical Package for Social Sciences;
WHO: World Health Organization

## Declarations

### Ethical approval and consent to participate

Ethical approval was obtained from the Institutional Review Board (IRB) of the College of Health Sciences at Addis Ababa University (protocol number: 042/20/SPH). Informed consent was obtained from all study participants.

### Consent for publication

Not applicable

### Author Contributions

WD, AW, WAA and WA conceptualized and designed the study. SG, WD and AW supervised the field data collection. WD and AK prepared the final dataset. WD, AK and AW analyzed and interpreted the data. WD drafted the manuscript. AW provided intellectual role in improving the manuscript. SG, AK, WAA and WD provided major roles in revising the manuscript. All authors read and approved the final manuscript.

## Acknowledgements

The authors are grateful to the research staff at the College of Health Sciences. The authors are also grateful to the data collectors and study participants for their time and contributing to the research.

