## Additional file 1 for "Knowledge and perceptions of COVID-19 among government employees in Ethiopia"

**Additional file 1**: List of institutions/organizations included in the survey with collected samples, June 2020

| No. | Government level<br>National Offices | Collected Sample (#) |
|---|---|---|
| 1 | Ministry of Health | 50 |
| 2 | Ministry of Education | 50 |
| 3 | Ministry of Science and Higher Education | 30 |
| 4 | Ministry of Finance | 50 |
| 5 | Ministry of Trade and Industry | 35 |
| 6 | Ministry of Labor and Social Affairs | 30 |
| 7 | Ministry of Women, Children and Youth | 27 |
| 8 | Ministry of Agriculture | 46 |
| 9 | Ministry of Transport | 34 |
| 10 | Ministry of Urban Development and Construction | 41 |
| 11 | Ministry of Innovation and Technology | 29 |
| 12 | Ministry of Culture and Tourism | 40 |
| 12 | Ministry of Water, Irrigation and Energy | 37 |
| 14 | Ministry of Revenues | 40 |
| 15 | National Bank of Ethiopia | 41 |
| 16 | Commercial Bank of Ethiopia (Addis Ababa Branch) | 44 |
| ***Subtotal*** | ***16*** | ***624*** |
| | **Addis Ababa City Administration Offices** | |
| 1 | Health Bureau | 40 |
| 2 | Education Bureau | 39 |
| 3 | Finance Bureau | 40 |
| 4 | Trade and Industry Bureau | 30 |
| 5 | Bureau of Labor and Social Affairs | 30 |
| 6 | Bureau of Women, Children and Youth | 28 |
| 7 | Bureau of Agriculture | 26 |
| 8 | Bureau of Transport | 38 |
| 9 | Bureau of Construction | 29 |
| 10 | Bureau of Culture and Tourism | 30 |
| 11 | Water and Sewerage Authority | 25 |
| 12 | Bureau of Revenues | 28 |
| 13 | Yeka Sub-City Administration Office | 39 |
| 14 | Lideta Sub-City Administration Office | 40 |
| 15 | Arada Sub-City Administration Office | 30 |
| 16 | Gulele Sub-City Administration Office | 40 |
| 17 | Bole Sub-City Administration Office | 40 |
| 18 | Addis Ketema Sub-city Administration Office | 38 |
| ***Subtotal*** | ***18*** | ***610*** |
| | **Oromia Regional State Offices** | |
| 1 | Health Bureau | 58 |
| 2 | Education Bureau | 18 |
| 3 | Finance Bureau | 30 |
| 4 | Bureau of Trade and Industry | 21 |
| 5 | Bureau of Labor and Social Affairs | 33 |

| | | |
|---|---|---|
| 6 | B. of Women, Children and Youth | 24 |
| 7 | Bureau of Agriculture | 33 |
| 8 | Bureau of Transport | 24 |
| 9 | Bureau of Urban Development and Construction | 26 |
| 10 | Bureau of Culture and Tourism | 21 |
| 11 | Bureau of Water, Irrigation and Energy | 21 |
| 12 | Bureau of Revenues | 30 |
| ***Subtotal*** | ***12*** | ***339*** |
| **Overall total** | **46** | **1,573** |
